# A quasi-experimental study to compare effectiveness of a breastfeeding arm sling with normal breastfeeding cross-cradle hold position

**DOI:** 10.1101/2023.01.24.23284943

**Authors:** Pornsri Disorntatiwat, Mary Steen, Sudjit Liblub

## Abstract

Breastfeeding has many benefits for the newborn and mother therefore, the World Health Organization guidelines recommend exclusive breastfeeding for the first 6 months, with continued breastfeeding for up to two years. However, exclusive breastfeeding rates in the first 6 months in Thailand were reported to be approximately, 14% in 2019. Research has highlighted that many mothers’ have a lack of belief in their ability to breastfeed, and some physical conditions, such as tiredness and difficulty continually holding their baby in a comfortable position. Additionally, first-time breastfeeding can contribute to mothers having difficulties breastfeeding during the early postnatal period. Therefore, the *Arm sling innovation* (device) has been designed to provide support and comfort whilst holding the newborn. This study compared the effectiveness of the breastfeeding arm sling innovation to support breastfeeding in cross-cradle hold position and normal cross-cradle hold position breastfeeding in first-time mothers. A quasi-experimental pretest-posttest research design was used to evaluate the effectiveness of breastfeeding before and after the intervention among first-time mothers in the postnatal unit, at Ramathibodi hospital, Thailand. A total of 46 postpartum mothers participated in the study. The results showed that the effectiveness of breastfeeding reported by mothers between using a normal cross-cradle hold position and using breastfeeding arm sling innovation was statistically significantly different (t = 4.32, P < 0.001) with helping to hold the baby securely without slipping (t=5.68, p<0.001) and mothers can continue to breastfeed (t=2.09, p <0.001). Majority of mothers were satisfied with the support of using the breastfeeding arm sling innovation design. The breastfeeding arm sling innovation contributes to the effectiveness of breastfeeding by assisting and supporting the mother and baby’s position to breastfeed more comfortably, thus assisting first-time mothers to feel comfortable, confident, and able to continue breastfeeding.

## Introduction

The benefits of breastfeeding for babies are well established as breast milk is naturally produced, safe, clean, provides complete nutrition, and contains antibodies that help protect against many common childhood illnesses [1]. Breastfeeding increases the bonding between a mother and her infant and also self-esteem for mothers from the experience [1]. Therefore, the World Health Organization has recommended that infants should be exclusively breastfed for the first six months of life and to continue breastfeeding for up to two years and beyond [2].

Despite the many benefits of breastfeeding, rates in Thailand remain low. World Health Organization guidelines and, also the most recent national report in Thailand have reported that the six months exclusive breastfeeding rate was approximately 14% in Thailand in 2019 [3]. According to the Ten Steps to Successful Breastfeeding, postpartum women should facilitate immediate initiation of breastfeeding, i.e., within the first hour after birth, and then attempt to breastfeed every 2–3 hours with a goal of eight sessions per 24-hour period to stimulate prolactin hormone to increase milk production [2,4]. However, there are a lot of factors that contribute to inadequate breastfeeding such as maternal discomfort which is frequently experienced by mothers during the breastfeeding period [5]. Previous research has found that many mothers have a lack of belief in their ability to breastfeed, and some physical conditions and tiredness can also contribute to them having difficulties breastfeeding during the early postnatal period [6]. Additionally, other problems can contribute to the first-time mothers’ inability to breastfeed their newborns, and some have reported feeling that they have insufficient milk and difficulties latching, thus leading to shortening the duration of breastfeeding [7]. It is important to note that breastfeeding experiences can vary considerably between first-time and previously breastfed mothers [8].

Positioning of the baby’s body and correct latching on of the baby is essential for good attachment and successful breastfeeding [9]. A cross-cradle hold position helps to guide the baby’s head and mouth while feeding and can help to avoid poor latching, which can cause nipple redness and/or cracking [10]. It has been reported that this position is preferred by new mothers who have little experience regarding breastfeeding or for babies who have not learned how to latch on the breast well [11]. Therefore, the midwife’s role involves helping mothers and providing support, such as guidance and counseling on practical concerns like breastfeeding positions and how to make mothers feel comfortable and thus promote longer duration of breastfeeding [12]. Signs that a baby is well attached during breastfeeding include holding the baby so that their face touches the mother’s breast, their head and body in alignment with the mother’s breast, their mouth wide open, and more of the areola is visible above the baby’s top lip than below the lower lip, as according to the four key signs for effective breastfeeding [2]. However, all these signs can lead to a mother feeling over-tired and can make her wrist and arm become uncomfortable and cause an aching sensation during breastfeeding.

Currently, there are several aids to help mothers breastfeed, for example, a functional nursing bra and a specifically designed supporting pillow; these aids can help mothers to continue breastfeeding. However, local new mothers have expressed how they find it difficult to continually hold their baby in a comfortable position and that their arm often aches during breastfeeding. The mothers’ experiences have been observed by the midwifery researchers and issues during breastfeeding such as, incorrect feeding position, extreme tiredness, and wrist pain have been identified whilst working in the postnatal unit at the local hospital (Ramathibodi Hospital). Therefore, there is a justification to find new ways to help and support first-time mothers to breastfeed comfortably and reduce the risk of wrist and arm pain as well as increase self-esteem for the mothers who choose to breastfeed.

## Definition of the methods of breastfeeding position

- Normal breastfeeding position is a normal cross-cradle hold position breastfeeding with a pillow support
- Breastfeeding arm sling innovation is the intervention to attach the mother’s arm whilst holding her baby to breastfeed in a cross-cradle hold position with a pillow support

## Aim and Objectives

- To compare the effectiveness of the breastfeeding arm sling innovation to support breastfeeding in cross-cradle hold position and normal cross-cradle hold position breastfeeding in first-time mothers.
- To evaluate the satisfaction with the breastfeeding arm sling innovation to support breastfeeding in cross-cradle hold position in first-time mothers
- The null hypothesis was that there is no difference in the mean scores for the effectiveness of breastfeeding between using breastfeeding arm sling innovation and breastfeeding in normal cross-cradle hold position.

## Materials and Methods

A quasi-experimental pretest-posttest research design was used to evaluate scores on the effectiveness of breastfeeding before and after using the breastfeeding arm sling innovation. This study design was chosen to undertake a comparison of the scores after an intervention to scores on the same measure in the same participants prior to the intervention [12]. Firstly, researchers designed and developed the breastfeeding arm sling innovation and conducted a pilot test of the innovation. Secondly, the researchers explained study information in a step-by-step approach guided by a research protocol and specific requirements of assessment to midwives (research assistants) for approximately, 30-60 minutes. The midwives demonstrated how they would undertake an assessment to confirm correct procedure.

Mothers who meet the inclusion criteria were invited to participate in the study. Mothers were informed about the purpose of the study and given an opportunity to discuss the study to make an informed choice to participate. Mothers were asked to complete a written consent form.

The experiment involved two stages (Fig 1). In the first stage, mothers were selected to provide 15 minutes of breastfeeding by using a cross-cradle hold position with the breastfeeding arm sling innovation or a normal cross-cradle hold position. As it is a clinical practice to wrap a baby (swaddle) to prevent hypothermia in the newborn in the postnatal unit at the study hospital [13-14], the babies in this experiment were wrapped in both groups (using a cross-cradle hold position with the breastfeeding arm sling innovation or a normal cross-cradle hold position), see Fig 1.

**Fig. 1.**
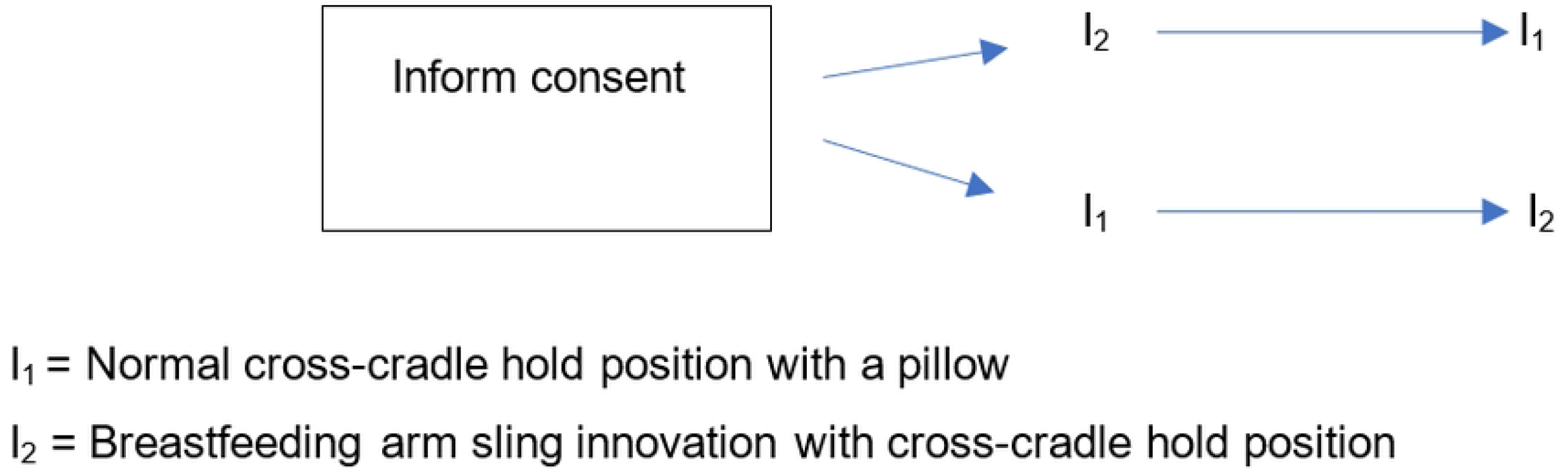
This is the Fig 1 Title. The experimental flowchart.

In the case of breastfeeding with a normal cross-cradle hold position, the midwives (research assistants) helped the mother to provide 15 minute-breastfeeding. The midwives (research assistants) observed and measured the effects of breastfeeding. In the case of breastfeeding with a cross-cradle hold position with the breastfeeding arm sling innovation, the midwives (research assistants) helped the mother to wear the sheath of the breastfeeding arm sling innovation and wrap the equipment around the baby. When the baby was latched on the midwives helped the mother to adhere the equipment for a firm hold. Following this, the mother provided 15-minute breastfeeding. The midwives observed and measured the effectiveness of breastfeeding. After breastfeeding, mothers were asked to complete the effectiveness of breastfeeding and satisfaction of using breastfeeding arm sling innovation questionnaires.

The following stage was undertaken three hours later, and 15-minute-breastfeeding was repeated (normal cross-cradle hold position or breastfeeding arm sling innovation with cross-cradle hold position) follow-up on the effectiveness of breastfeeding and satisfaction levels outcomes were assessed.

### Participants and setting

This study was conducted in the postnatal unit, at Ramathibodi hospital, Thailand. Two hours after giving birth, mothers were transferred to the postnatal unit and provided with care and support. It is normal practice for healthy mothers and their newborn babies to be discharged home from the postnatal unit within 48-72 hours. In this unit, assistance with breastfeeding during the postpartum period is provided by nurse-midwives. Research assistants, thus, are nurse-midwives who are healthcare providers in the postnatal unit, having knowledge of assessing and observing breastfeeding behaviour. Breastfeeding assistance can vary according to the nurse-midwife that provides the care. Therefore, during the undertaking of this study, the nurse-midwife who supported a mother to breastfeed continued to be the same person, in order to control for any breastfeeding effect.

### The structured questionnaire

The effectiveness of breastfeeding was recorded by a questionnaire for participants and midwife assessors that was modified from the four-key points (The Breastfeed Observation), which aligned with the key signs to effective breastfeeding. Questions were related to the four key points of the baby’s position: baby’s head and body in line, baby held close to mother’s body, baby faces the breast with nose opposite the nipple, baby’s whole body supported [2].

Additionally, observations relating to good attachment: more areola seen above baby’s top lip, baby’s mouth open wide, lower lop turned outwards and baby’s chin touches breast were considered. The participants self-reported these effective breastfeeding signs, and the midwives observed these signs. Both questionnaires included five items that utilised a ranked 5-point Likert-type scale to assess the effectiveness of breastfeeding outcomes.

The breastfeeding arm sling innovation satisfaction questionnaire was developed by the researchers and adapted from the diffusion of innovation model, which demonstrates innovation is generally adopted when four relative advantages are considered i.e., compatibility, complexity, trial-ability, and observability [15]. The questionnaire includes 14 items ranked on a 5-point scale from lowest satisfaction to the highest level of satisfaction. The face and content validity were tested through consultation with a panel of three experts, including maternal-newborn and midwifery experienced researchers from the Ramathibodi School of Nursing, Mahidol University. The structure of the questionnaire also included open-ended questions and sought qualitative information regarding using the breastfeeding arm sling innovation to explore what participants views were and report their thoughts in their own words [16].

### Sample size calculation

The sample size was calculated based on a pilot study involving 5 mothers that were undertaken in the same setting to test the study feasibility (SD. = 1.2). According to the formula to calculate the sample size, considering an alpha error of 0.05, Delta 0.5, and a test power of 80%, under one tail hypothesis, the sample size of 46 mothers was required to participate [17].

### Data collection

Recruitment and data collection were undertaken in the postnatal unit on a daily basis, by a research assistant, who recruited participants and implemented the assignment of the intervention. A midwife (research assistant) supported and advised a mother how to breastfeed using the cross-cradle holding position and how to use the breastfeeding arm sling innovation. The mothers who met the inclusion criteria were asked to assess their breastfeeding effectiveness before the intervention. After the breastfeeding intervention (15 minutes), the mothers were asked to assess their breastfeeding effectiveness and measure self-reported satisfaction. The midwives (research assistants) observed and completed the effectiveness of the breastfeeding questionnaire in both normal breastfeeding and during the intervention.

### Intervention

The breastfeeding arm sling innovation has been designed and tailored by using spandex for the sleeves, filled with elastic and attached with 6 cm leather, see Fig. 2. The spandex used for both arm wear is 5.0 cm. apart and has two straps that are 63 × 7.5 cm. Both ends of the spandex are attached to Velcro straps in two positions to allow adjustment of the size according to the baby’s size.

**Fig 2.**
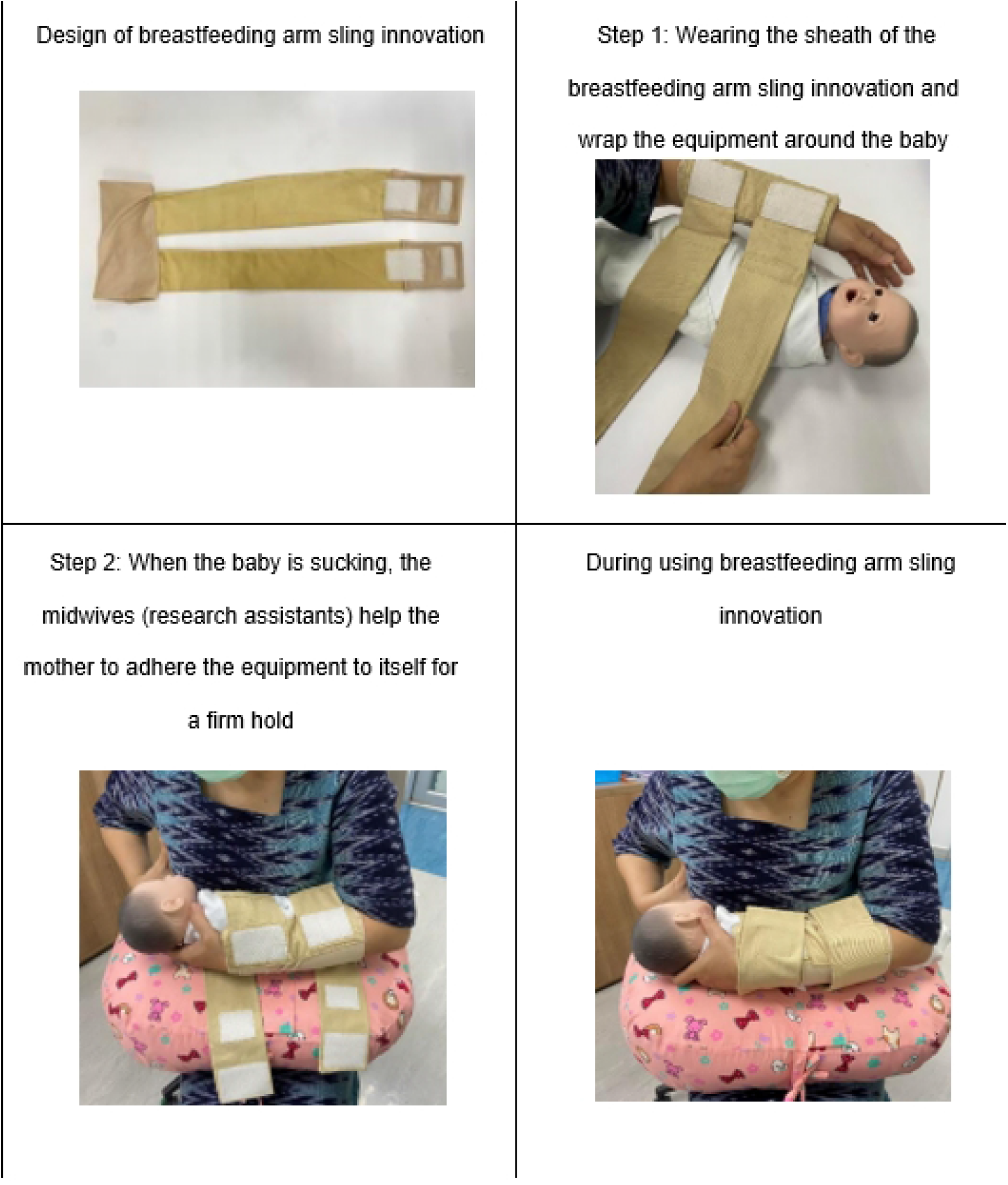
This is the Fig 2 Title. Breastfeeding arm sling innovation.

In the first step of applying the support equipment, the midwives (research assistants) helped the mother to wear the sheath of the breastfeeding arm sling innovation and wrap the equipment around the baby. When the baby was latched on and feeding, the midwives (research assistants) helped the mother to adhere the equipment for a firm hold, so that the mother’s arm touched closely the baby’s back to support breastfeeding. This technique helped the mother’s arm to touch the baby’s back, therefore enabling her to hold the newborn baby closer. An advantage of this approach is that it prevents fatigue of the wrists and shoulders of the mothers during breastfeeding.

As this research is in the experimental stage, a helper was required to support how to apply the breastfeeding arm sling innovation. This initial help to use the breastfeeding arm sling innovation showed mothers how to apply and use the breastfeeding arm sling by themselves for further breastfeeding (Fig 2).

### Statistical analysis

All data analysis was performed using the IBM SPSS Statistics 27.0.1.0 software package [18]. Data were expressed as means and standard deviations (SD). A comparison of the means of the two groups within subjects was evaluated for significance using paired t-tests. A p-value of less than 0.05 was considered significant.

## Ethical considerations

The study was approved by the Ethics Committee on Human Subjects of the Faculty of Medicine at Ramathibodi Hospital, Mahidol University, Thailand (IRB COA. MURA2021/949), in line with the Human Research Protection such as Declaration of Helsinki, The Belmont Report, and the International Conference on Harmonization in Good Clinical Practice (ICH-GCP).

## Results

### Participants’ demographic characteristics

A total of 46 postpartum women participated in the study ranging in age from 19 to 40 years with a mean age of 29 years (SD = 5.65). Gestational age ranged from 37-41 weeks (mean gestational age 38 weeks, SD = 1.09 weeks). Mothers who participated in the study were post-birth on their first day n = 18 (54.5 %), second day n= 9 (27.3 %), and third day n=6 (18.2%). The babies of the participants had a mean weight of 3151 grams.

### Differences in the effectiveness of breastfeeding observed by mothers between groups

The results of paired t-tests showed that the effectiveness of breastfeeding reported by mothers between using a normal cross-cradle hold position and using a cross-cradle hold position with breastfeeding arm sling innovation was statistically significantly different (t = 4.32, P < 0.001). The difference in total scores of using breastfeeding arm sling innovation and normal breastfeeding was statistically significant (hold the baby securely without slipping: t=5.68, p<0.001; feeling no pain at the nipples while the baby is suckling: t=4.76, p <0.001; baby can continue to suck: (t=2.09, p <0.001); baby held close to mother’s body: t=5.76, p <0.001; No pain felt at the breast during sitting to breastfeed continuously: t=4.24, p <0.001), as presented in table 1.

**Table 1.** Comparison of the effectiveness of breastfeeding between using normal cross-cradle hold position and using cross-cradle hold position with breastfeeding arm sling innovation by mothers.

| Effectiveness of breastfeeding | Using<br>breastfeeding<br>arm sling<br>innovation<br>equipment |  | Normal<br>breastfeeding |  | t | p-value |
| --- | --- | --- | --- | --- | --- | --- |
|  | MEAN | SD | MEAN | SD |  |  |
| 1. Hold the baby tightly without slipping | 3.24 | 0.67 | 2.20 | 1.02 | 5.68 | <0.001 |
| 2. The feeling of no pain at the nipples while the baby is suckling | 2.63 | 0.74 | 2.04 | 0.89 | 4.76 | <0.001 |
| 3. Baby can continue to suck | 3.02 | 0.86 | 2.33 | 0.97 | 2.09 | <0.001 |
| 4. Baby held close to mother's body | 3.28 | 0.78 | 2.24 | 1.08 | 5.76 | <0.001 |
| 5. No pain felt at the breast during sitting to breastfeed continuously | 2.65 | 0.77 | 1.96 | 0.89 | 4.24 | <0.001 |
| Total | 2.97 | 0.60 | 2.15 | 0.80 | 4.32 | <0.001 |

### Differences in the effectiveness of breastfeeding observed by midwives between groups

Table 2 shows the effectiveness of breastfeeding observed by the midwife and indicates a significant difference in the effects of signs of baby attached during breastfeeding between normal breastfeeding and using breastfeeding arm sling innovation in all outcomes. Compared with normal breastfeeding, using breastfeeding arm sling innovation increased significantly as greater areola seen above the baby’s top lip (t = 6.03, p <0.001); baby’s chin touches breast (t = 6.26, p <0.001); baby’s head and body in line (t = 7.46, p <0.001); baby’s mouth open wide (t = 6.49, p <0.001); baby’s cheeks do not dimple during breastfeed (t = 7.29, p <0.001).

**Table 2.** **Comparison of the effectiveness of breastfeeding between using normal cross-cradle hold position and using cross-cradle hold position with breastfeeding arm sling innovation by midwife (researcher assistance)**

| Effectiveness of breastfeeding | Using breastfeeding arm sling innovation equipment |  | Normal breastfeeding |  | t | p-value |
| --- | --- | --- | --- | --- | --- | --- |
|  | MEAN | SD | MEAN | SD |  |  |
| 1. More areola seen above baby's top lip | 2.93 | 0.77 | 2.22 | 0.59 | 6.03 | <0.001 |
| 2. Baby's chin touches breast | 3.08 | 0.66 | 2.34 | 0.71 | 6.26 | <0.001 |
| 3. Baby's head and body in line | 3.24 | 0.77 | 2.04 | 0.87 | 7.46 | <0.001 |
| 4. Baby's mouth open wide | 3.24 | 0.82 | 2.37 | 0.77 | 6.49 | <0.001 |
| 5. Baby's cheeks do not dimple during breastfeeding | 3.30 | 0.76 | 2.48 | 0.72 | 7.29 | <0.001 |
| Total | 3.16 | 0.64 | 2.29 | 0.59 | 8.93 | <0.001 |

### Mothers’ satisfaction using breastfeeding arm sling innovation

The majority of the mothers were satisfied with the suitability of using the breastfeeding arm sling innovation design (52.9%, size and length 44.1% and fabric used 41.2%). While half (50%) of the mothers were satisfied with the appearance of the breastfeeding arm sling innovation, (41.2%) reported that they were very satisfied with the non-slippage. Meanwhile, satisfied level of the features of easy to use (41.2%), ease of disassembly (41.2%), and ease of cleaning (38.2%) were reported by the mothers (Table 3).

**Table 3.** Mothers’ satisfaction using breastfeeding arm sling innovation.

| How satisfied are you with using the breastfeeding arm sling innovation | Satisfaction levels (%) |  |  |  |  |
| --- | --- | --- | --- | --- | --- |
|  | Very Satisfied | Satisfied | Neutral | Unsatisfied | Very Unsatisfied |
| **Easy-to-use features** |  |  |  |  |  |
| 1) Easy to use | 17.6 | 41.2 | 35.3 | 2.9 | 2.9 |
| 2)Non slippage | 41.2 | 41.2 | 17.6 |  |  |
| 3)Easy to disassemble | 5.9 | 41.2 | 41.2 | 11.8 |  |
| 4)Easy to clean | 38.2 | 38.2 | 23.5 |  |  |
| **Suitability of equipment** |  |  |  |  |  |
| 5)Design | 14.7 | 52.9 | 29.4 | 2.9 |  |
| 6) Size and length | 26.5 | 44.1 | 26.5 | 2.9 |  |
| 7) Beauty | 11.8 | 50.0 | 35.3 | 2.9 |  |
| 8) Fabric used | 32.4 | 41.2 | 23.5 | 2.9 |  |
| **Trial-ability** |  |  |  |  |  |
| 9)The ability to hold a baby to<br>breastfeed | 35.3 | 41.2 | 17.6 | 5.9 |  |
| 10)Ease of carrying a baby | 35.3 | 41.2 | 17.2 | 5.9 |  |
| 11)Pain in the arm while holding<br>the baby | 35.3 | 29.4 | 23.5 | 11.8 |  |
| 12)Tension in the wrist while<br>holding the baby | 20.6 | 44.1 | 26.5 | 8.8 |  |
| **Observability** |  |  |  |  |  |
| 13)Satisfaction with the use of the<br>innovation for breastfeeding | 32.4 | 38.2 | 23.5 | 5.9 |  |
| 14)Feelings of Confidence in<br>Breastfeeding | 38.2 | 35.3 | 20.6 | 5.9 |  |

Regarding the trial-ability, (44.1%) of mothers reported the breastfeeding arm sling innovation helped to reduce tension in the wrist while holding the baby. Mothers reported that they were satisfied with their ability to hold their baby to breastfeed (41.2%) and ease of carrying baby (41.2%). Overall, (32.4%) of mothers felt very satisfied with the use of breastfeeding arm sling innovation and (38.2%) reported feeling confident in breastfeeding.

## Discussion

The development of innovations model is a new concept in the nursing and midwifery process for developing, improving, and adapting products or procedures to significantly benefit outcomes [10]. Results of the present study show the clinical usefulness of the breastfeeding arm sling innovation in supporting breastfeeding. The findings from this current study showed that mothers and midwives reported the effectiveness of breastfeeding between using a normal cross-cradle hold position and using a cross-cradle hold position with breastfeeding arm sling innovation was statistically significantly different in all-term supports (t= 4.32, p <0.001) included holding the baby so that their face touched the mother’s breast, their head and body was in alignment with their mother’s breast, mouth wide open, and more of the areola was visible above the baby’s top lip than below the lower lip. These findings are in accordance with Rahim et al. study, who emphasised success of the breastfeeding process depends on the accuracy of the position and attachment of the baby to the mother’s breast and the ability of the baby to suck [19].

For a good attachment and effective breastfeeding, the baby’s body positioning and latching technique are crucial [9]. Improper latch and inappropriate infant positioning can cause nipple pain and trauma [20]. Results from this current study demonstrate that the breastfeeding arm sling innovation helped mothers to breastfeed and decreased the feeling of pain at the nipples while the baby was latched on and feeding. This significant improvement in comfort helped to encourage continued breastfeeding.

The use of an innovative breastfeeding arm sling helped the mothers who participated in this study to hold the baby’s body without any slipping during breastfeeding so that the baby’s mouth and head were in a position in alignment with the nipple. Correct positioning and attachment of newborns during breastfeeding was one of the recommendations by Kent et al. [21], to help reduce nipple pain, increase the duration of breastfeeding, and reduce breastfeeding problems.

Breastfeeding can be challenging for first-time mothers who have little experience of breastfeeding and newborns who have not yet learned how to properly latch on to the breast [11]. The current study’s results show that the breastfeeding arm sling innovation helped mothers feel less pain in the arm while holding the baby and sitting to breastfeed continuously. The study’s findings are consistent with previous research demonstrating that innovations can address patient demands or help solve problems, leading to better outcomes for patients [10]. The first-time mothers in this study valued the breastfeeding arm sling innovation and felt very satisfied as it helped them feel comfortable during breastfeeding. This may be due to the breastfeeding arm sling innovation fitting the mother’s arm to promote a correct position and enabling her to touch the newborn baby’s back so that she can hold her baby closer during breastfeeding. Given the appropriate posture during breastfeeding, the breastfeeding arm sling innovation is considered acceptable among first-time mothers, with most mothers using it and reporting more confidence in breastfeeding.

The breastfeeding arm sling innovation was reported to have very satisfactory features due to its non-slipping benefit. With the features of easy to use and disassemble, however, this breastfeeding arm sling innovation required assistance to help mothers apply the equipment, wrapping it around the baby in order to support the correct position when the baby was breastfeeding. Self-application needs to be factored in when considering the development of the breastfeeding arm sling innovation in the future to help mothers breastfeed independently.

A strength of this study is that mothers were very willing to participate in the study and this is also reflected in the high levels of satisfaction reported. Midwife assessors had preparatory training to assist mothers who agreed to participate. However, the assessors were known to the mothers, and this may have influenced their views and experiences positively or negatively as continuity of care is beneficial but may also have an impact on outcomes. The quasi-experimental study design allowed the participants to be their own control group, but findings are self-reported and must be viewed cautiously. The research study used a non-randomised design, therefore, other confounding factors not controlled for may have influenced the findings and therefore this limits the inference of the research results. Another limitation concerning this study was that it was limited to only first-time mothers, which makes it difficult to generalise the findings to other groups of childbearing women. Since the breastfeeding arm sling innovation in the present study applies to term newborn babies, future development would need to consider investigation as to whether preterm and differences in the baby weights would achieve similar results as reported in the present study.

## Conclusions

The development of innovations model in nursing is a new concept in the nursing process for developing, improving, or adapting products or procedures to significantly benefit outcomes and respond directly to patient needs. The breastfeeding arm sling breastfeeding innovation contributes to the effectiveness of breastfeeding by assisting and supporting the mother and baby’s position to breastfeed more comfortably. In conclusion, this study provides emerging evidence that the breastfeeding arm sling assisted first-time mothers to feel comfortable, confident, and helped them to continue breastfeeding. However, further research is required to confirm or refute these initial findings.

## Data Availability

All data without restrctions will be available

## Acknowledgments

The authors would like to gratefully thank the Faculty of Medicine, Ramathibodi Hospital, Mahidol University for their support grant and all the participants who took part in the present study.

